# Type 2 diabetes as a determinant of Parkinson’s disease risk and progression

**DOI:** 10.1101/2020.11.12.20230474

**Authors:** Harneek Chohan, Konstantin Senkevich, Radhika K Patel, Jonathan P Bestwick, Benjamin M Jacobs, Sara Bandres Ciga, Ziv Gan-Or, Alastair J Noyce

## Abstract

**Objective:** To investigate type 2 diabetes mellitus (T2DM) as a determinant of Parkinson’s disease (PD) through a meta-analysis of observational and genetic summary data.

**Methods:** A systematic review and meta-analysis of observational studies was undertaken by searching six databases. We selected the highest quality studies investigating the association of T2DM with PD risk and progression. We then used Mendelian randomization (MR) to investigate causal effects of genetic liability towards T2DM on PD risk and progression, using summary data derived from genome-wide association studies.

**Results:** In the observational part of the study, nine studies were included in the risk meta-analysis and four studies were included in the progression meta-analysis. Pooled effect estimates revealed that T2DM was associated with an increased risk of PD (OR 1.21, 95% CI 1.07-1.36), and there was some evidence that T2DM was associated with faster progression of motor symptoms (SMD 0.55, 95% CI 0.39-0.72) and cognitive decline (SMD −0.92, 95% CI −1.50 – −0.34). Using MR we found supportive evidence for a causal effect of diabetes on PD risk (IVW OR 1.08, 95% CI 1.02-1.14; p=0.010) and some evidence of an effect on motor progression (IVW OR 1.10, 95% CI 1.01-1.20; p=0.032), but not for cognitive progression.

**Conclusion:** Using meta-analysis of traditional observational studies and genetic data, we observed convincing evidence for an effect of T2DM on PD risk, and new evidence to support a role in PD progression. Treatment of diabetes may be an effective strategy to prevent or slow progression of PD.

## INTRODUCTION

Type 2 diabetes (T2DM) and Parkinson’s disease (PD) are prevalent diseases which affect an aging population. Emerging evidence suggests biological links between the two. Both are characterised by aberrant protein accumulation, lysosomal and mitochondrial dysfunction, and chronic systemic inflammation.^1,2^ Insulin resistance is a hallmark of T2DM and may be an important contributing factor to PD too.^3^

Previous systematic reviews and meta-analyses have explored the link between diabetes and the risk of PD, but the results are conflicting. For instance, pooled effect estimates from case-control studies suggest that diabetes has a negative association with PD risk,^4,5,6^ whilst meta-analyses that focus on prospective cohort studies suggest an increased risk of PD in patients with diabetes.^5,7^ Importantly, these analyses included patients with diabetes in general, rather than T2DM specifically, and have not considered the effect that T2DM has on progression of PD.

The association between T2DM and risk of PD has also not been explored thoroughly using modern causal methods. Mendelian randomization (MR) is a method in genetic epidemiology that can be used to follow-up observational associations for evidence of true causal effects.^8^ Genetic variants are distributed randomly at birth, meaning the determinant (in this case T2DM) is not affected by the presence of the outcome (here PD) and confounding factors are also randomly distributed. The current study combines meta-analysis of observational data with meta-analysis of genetic data (MR) to evaluate the effect that T2DM has on the risk of developing PD, and on motor and cognitive progression in patients with PD.

## METHODS

### Observational study data

#### Literature searches

The Meta-analyses of Observational Studies in Epidemiology (MOOSE) guidelines were followed to conduct the literature search. Various electronic databases were used, including PubMed, Web of Science, Scopus, and Ovid, which incorporates the Embase and Medline databases. Additionally, the preprint electronic servers Biorxiv and Medrxiv were used for a more comprehensive search of recent literature, which has not been peer-reviewed. The searches took place between 2nd June 2020 until 6th June 2020; no filters were applied during the search.

The terms used for searches were “Type 2 Diabetes” AND “Parkinson’s disease” OR “Type 2 Diabetes” AND “progression of Parkinson’s disease” OR “Type 2 diabetes” AND “Parkinson’s disease risk”. The reference lists of included articles were hand-searched to ensure the inclusion of any articles which may have been missed by the electronic searches. Independent literature searches were conducted by two authors (HC and RP).

#### Inclusion Criteria

The inclusion of studies for the main meta-analysis consisted of (1) observational studies that investigated preceding T2DM specifically and its effect on the risk of PD and (2) studies analysing how diabetes was associated with progression of PD. Observational studies of the association between diabetes (in general) and risk of PD were analysed separately.

#### Exclusion Criteria

Articles which were not observational studies were excluded, such as reviews, short surveys, books, and letters that reported no new data (see Figure 1). Studies reporting on drug-induced Parkinsonism and any articles not written in English were excluded.

**Figure 1.**
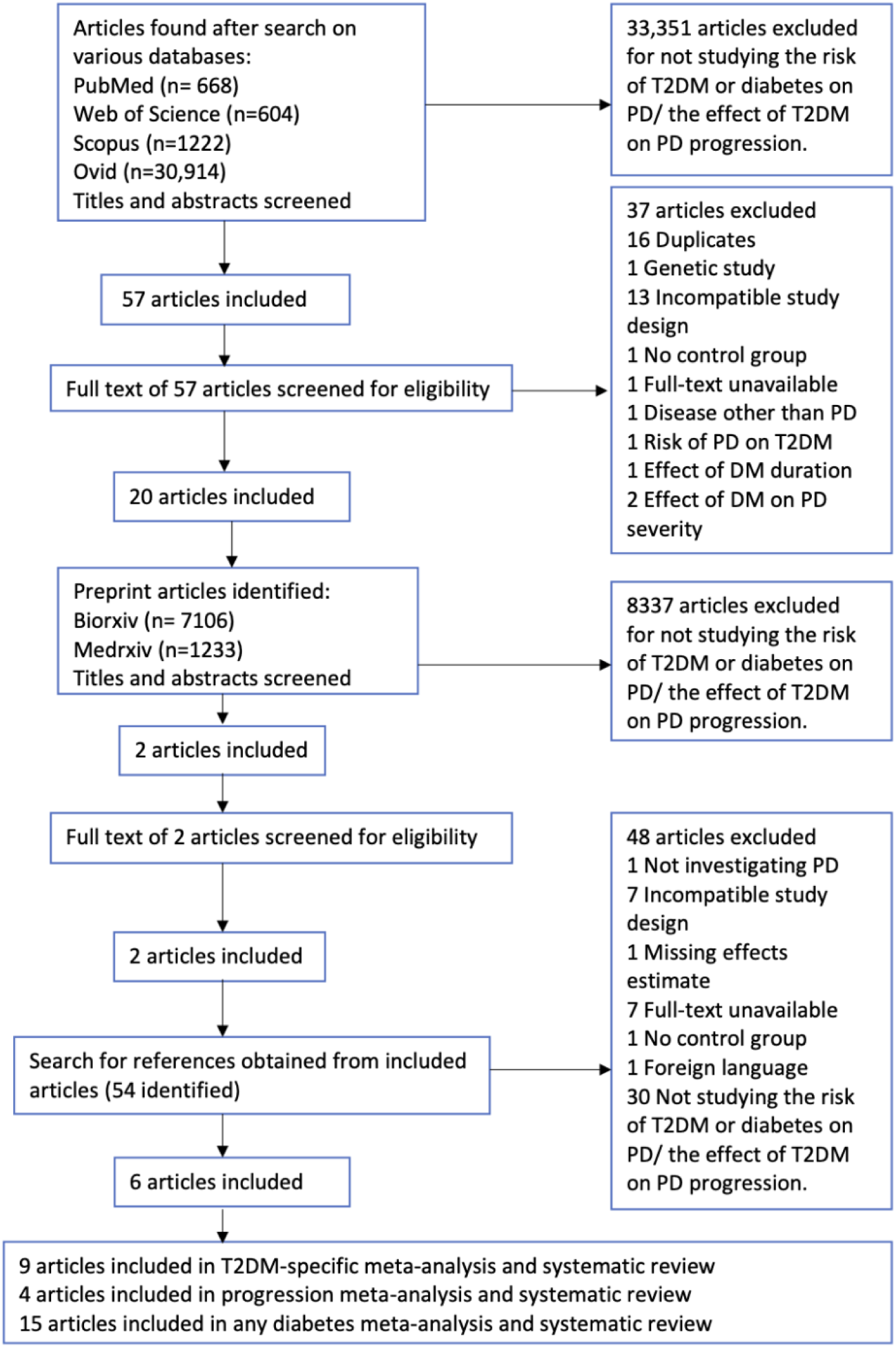
PRISMA flowchart used for the systematic review process.

#### Data Extraction

The Preferred Reporting Items for Systematic Reviews and Meta-Analyses (PRISMA) guidelines were followed for data extraction.^9^ The data extracted from the articles included the study design, the effect estimates and the 95% confidence intervals (CI), the length of follow-up, and any adjustments made for confounding factors. For studies reporting data for males and females separately, the effect estimates for both sexes combined were used. The Newcastle Ottawa Scale (NOS) was used to assess the quality of the studies included.^10^

#### Statistical Analysis

Effect estimates from individual studies of PD risk, including odds ratios (ORs), relative risks (RRs), hazard ratios (HRs) and incidence rate ratio (IRR) were used to calculate a combined effect estimate for the association between T2DM and risk of PD using standard meta-analysis methods. For PD progression, we used standard motor scales (such as part III of the UPDRS or Hoehn and Yahr stage) and cognitive scales (such as the MoCA or the MMSE). The mean progression per year was calculated to generate a standardised mean difference (SMD) for each study on a 1-year scale. For studies providing a ratio effect estimate, the ratio was also converted into a SMD.^11^ For all meta-analyses, the effect estimates were pooled using fixed-effects and weighted by inverse variance, before being assessed for heterogeneity using the I2 statistic. Where there was evidence for heterogeneity (I2 >50% and p-value <0.05) the meta-analysis was re-run using a random-effects model to combine effect estimates. As a result of these steps, a random-effects model was used to combine estimates for risk and a fixed-effects model was used for progression. To assess the heterogeneity further, and to check for bias causing the differences in estimates in the studies investigating risk, the studies were separated by study design and time of patient enrolment for sub-analyses (patients enrolled when the exposure (T2DM) is observed, patients enrolled before T2DM is observed and studies enrolling patients before and after exposure observation). Where there was evidence for publication bias, a trim and fill analysis was performed. Meta-regression was performed to investigate the effect of gender and age on effect estimates. Stata version 16.0 was used to perform these analyses.

### Mendelian randomization

#### Exposure instrument

A recent multi-ethnic meta-analysis of genome wide association studies (GWAS) on T2DM was selected as exposure to construct a genetic instrument for liability towards T2DM.^12^ To avoid bias due to population differences, we selected only GWAS significant SNPs (p-value < 5×10^−8^) from participants of European ancestry (total 425 SNPs; N cases = 148,726, N controls = 965,732). We performed clumping with standard parameters (clumping window of 10,000 kb, R^2^ cut-off 0.001) to exclude variants in linkage disequilibrium (LD). The proportion of variability explained by genetic variants (R^2^) and the strength of association with the exposure (F-statistic) were calculated using standard methods.^13,14^

#### Outcome data

For PD risk as an outcome, we used the summary statistics from the latest PD GWAS meta-analysis.^15^ Since participants from UK Biobank (UKB) data was present in both the T2DM GWAS and PD GWAS, we used a subset of the PD GWAS summary statistics which excluded UKB and 23andMe participants (N cases = 15,056, N controls = 12,637).

For PD progression, we sought similar data to that from the observational study meta-analysis (i.e. UPDRS part 3, MMSE and MoCA scores), derived from GWASs of these PD progression traits as outcomes.^16^ Sample sizes for PD progression studies were calculated as mean across all SNPs in GWAS summary statistics. Based on the following sample sizes (PD risk, N= 27,693; UPDRS part 3, N= 1398; MMSE, N = 1329; MoCA, N= 1000), we calculated power to detect an OR of 1.2 for each outcome (https://sb452.shinyapps.io/power/).^17^

#### Statistical Analysis

MR analyses were performed using two-sample MR package in R.^18,19^ Steiger filtering was applied to exclude SNPs that explained more variance in outcome than in exposure.^20^ We calculated pooled causal estimates using inverse-variance weighted method (IVW).^19^ A variety of sensitivity analyses were performed using MR-Egger and weighted median methods. Mendelian Randomization Pleiotropy RESidual Sum and Outlier method (MR-PRESSO) was performed to detect pleiotropic outliers and adjust where necessary.^21^ Cochran’s Q test in the IVW and MR-Egger methods was performed to assess for evidence of heterogeneity.^22^

## RESULTS

### Observational study data

The literature search produced 33,408 articles of which 28 were eligible for the meta-analyses. Details regarding the exclusion of articles can be found in Figure 1. Details of the included studies can be found in Supplementary tables 1-3. Quality assessment using the NOS (Supplementary tables 4-6) revealed that studies included in the risk and progression meta-analyses were generally of good quality.

#### T2DM and Risk of PD

Of the nine studies that defined T2DM as the exposure, seven were cohort studies, and two were case-control studies. The overall effect estimate was 1.21 (95% CI 1.07-1.36; see Figure 2). When considered separately, cohort studies (OR 1.29, 95% CI 1.19-1.40) provided strong evidence for T2DM being associated with higher PD risk, but there was an inverse association between T2DM and PD in case-control studies (OR 0.51, 95% CI 0.30-0.87). The funnel plot combining all nine studies was visually asymmetric suggesting the possibility of publication bias (Supplementary figure 1), but the accompanying p-value was 0.145.

**Figure 2.**
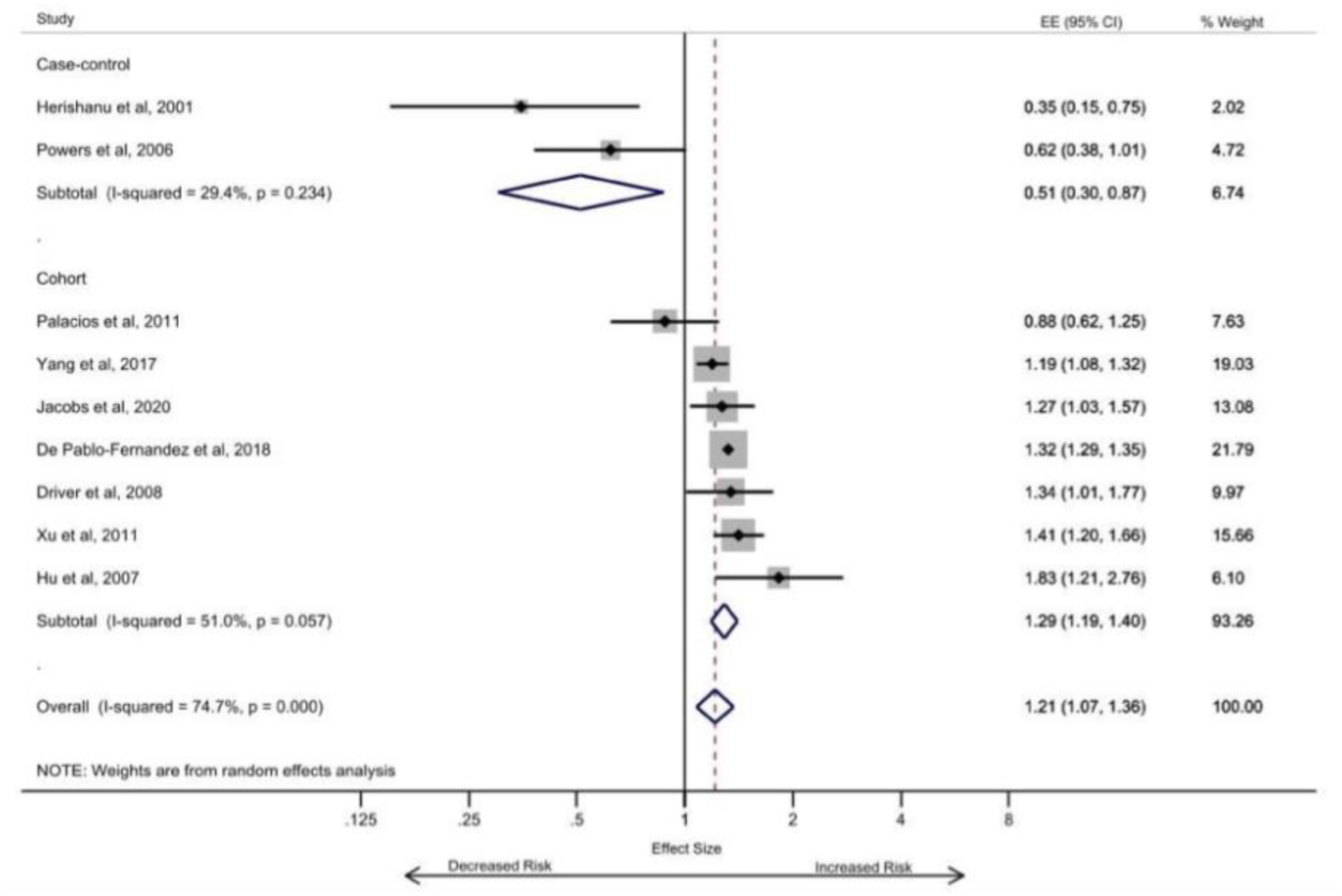
Type 2 diabetes increases the risk of developing Parkinson’s disease (T2DM-specific analysis).

We explored whether there was a change in PD risk estimate according to the average age of participants in cohort studies (Supplementary figure 2), but there was no evidence to support this (p=0.540). Age was not investigated in the case-control studies due to insufficient observations. Similarly, there was no convincing evidence that the ratio of males and females accounted for differential study effects (p=0.165). We then considered the cohort studies separately depending on when enrolment to the study occurred relative to the onset of the exposure (T2DM), under the assumption that the effect of survival bias in those with exposure prior to enrolment (i.e. unobserved) would be greatest.^23^ In one cohort study, the onset of T2DM was prior to the period of observation (OR 0.88, 95% CI 0.62-1.25), two of the cohort studies enrolled incident cases of T2DM (pooled OR 1.27, 95% CI 1.15-1.40), whilst the remaining four cohort studies used a combination of methods to ascertain the T2DM (pooled OR 1.38, 95% CI 1.24-1.55) (Supplementary figure 3).

We repeated the risk meta-analysis using the studies that failed to specify T2DM as the exposure and included patients with *any* diabetes. These studies were generally lower in quality (Supplementary table 5). The combined effect estimate for all studies was 1.08 (95% CI 0.92-1.27), giving an overall null effect for the association between diabetes and risk of PD (Supplementary figure 4). However, there was a clear divergence in the pooled effect from case-control studies (OR 0.79, 95% CI 0.54-1.17) and cohort studies (OR 1.28, 95% CI 1.05-1.56), which was greater than that observed in the studies that focused on T2DM specifically. There was also evidence of publication bias in the combined analysis of case-control and cohort studies (p=0.01; Supplementary figure 5). The trim and fill sensitivity analysis (Supplementary figure 6) led to a small increase in the pooled effect estimate to 1.12 (95% CI 0.95-1.31). Meta-regression revealed no evidence that the ratio of males to females was a determinant of the differential association between diabetes and PD risk across studies (p-value 0.542). In case-control studies, as the average age of participants rose, the association with PD risk changed from negative to positive, but there was only weak evidence to suggest that this was anything other than a chance finding (p=0.043; Supplementary figure 7). There was no similar change in effect by participant age in the cohort studies (p=0.268; Supplementary figure 8) and when we pooled case-control studies that specified T2DM with those that included any diabetes, there was no convincing effect of age (p=0.053; Supplementary figure 9).

#### Diabetes and Progression of PD

Pooling data from three studies provided evidence for diabetes being associated with a faster progression in the severity of motor symptoms in PD patients (Figure 3). The overall annual SMD was 0.55 (95% CI 0.39-0.72), suggesting that diabetes was associated with more rapid progression of motor features in PD. Pooling data from two studies provided evidence for diabetes also being associated with a faster cognitive decline in PD patients (Figure 4). The overall annual SMD for cognitive change was −0.92 (95% CI −1.50 – −0.34).

**Figure 3.**
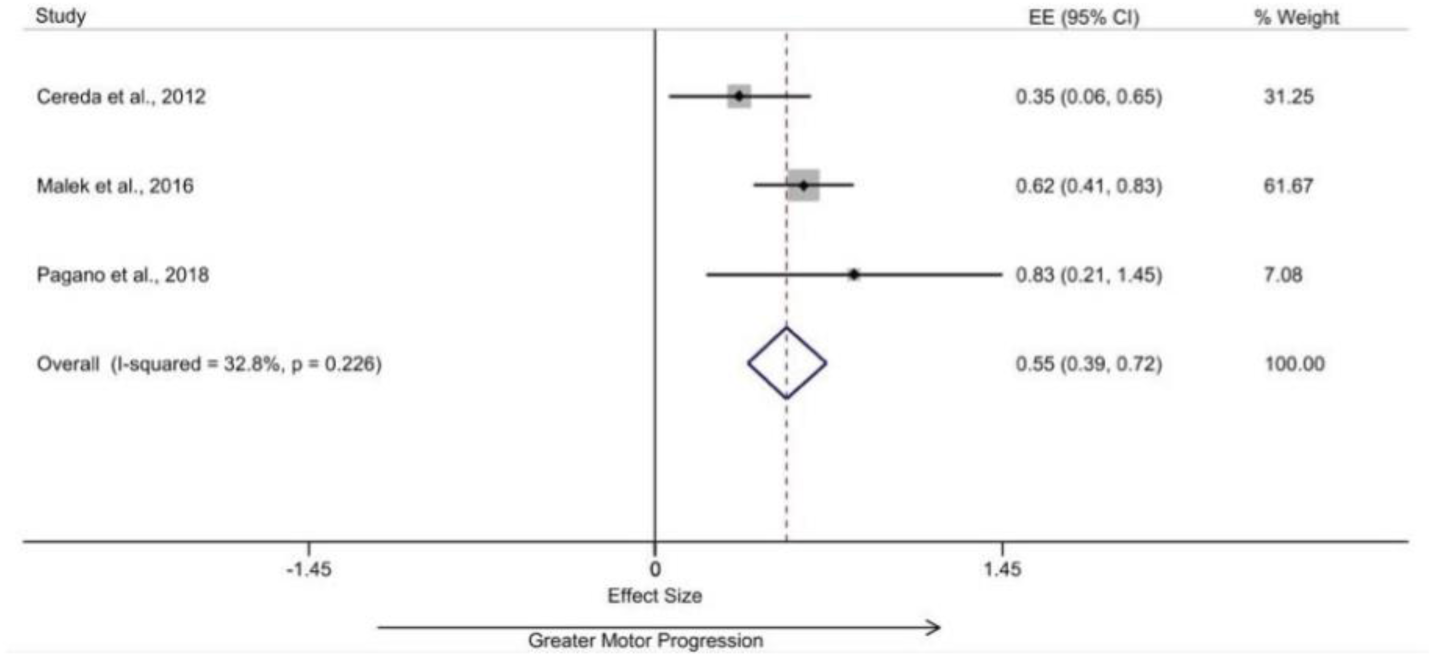
Diabetes results in greater motor progression in patients with Parkinson’s disease.

**Figure 4.**
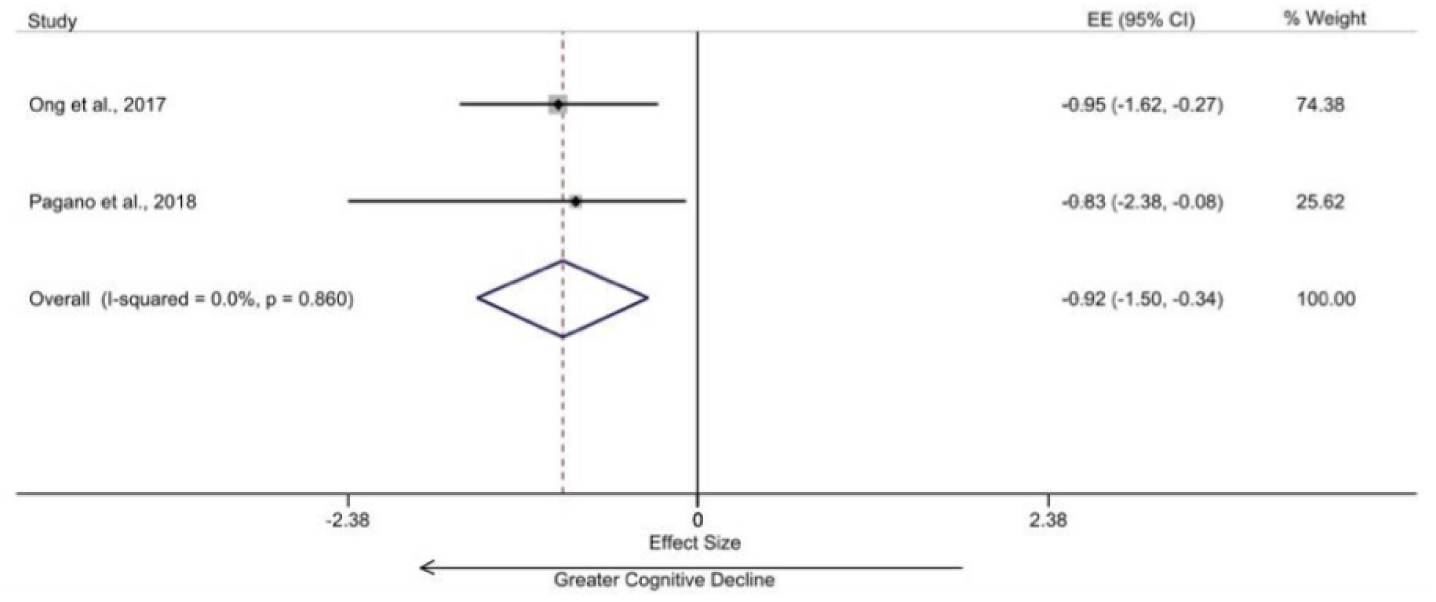
Diabetes results in greater cognitive decline in patients with Parkinson’s disease.

### Mendelian randomization

Separately, we performed MR to assess evidence for a causal effect of T2DM on PD risk and progression. After clumping, the total number of independent SNPs that comprised the exposure instrument (liability towards diabetes) was 191. The instrument had a R^2^ = 1.5% and the F-statistic was 51.2, with an F statistic of 10 generally considered to indicate an instrument of sufficient strength for causal analysis. We found evidence for a causal effect of liability towards T2DM on PD risk (IVW OR=1.08, 95% CI 1.02-1.14; p=0.010; Table 1; Supplementary Table 7; Supplementary Figures 10-12).

**Table 1.** MR analysis between exposure (T2DM) and outcomes (PD-risk and progression).

| Outcome | N, SNPs included | Power | Inverse variance weighted |  | MR Egger |  |
| --- | --- | --- | --- | --- | --- | --- |
|  |  |  | OR (95% CI) | p-value | OR (95% CI) | p-value |
| PD risk | 185 | 38.00% | 1.08 (1.02-1.14) | 0.010 | 1.05 (0.93-1.17) | 0.427 |
| Continuous PD progression traits |  |  |  |  |  |  |
| UPDRS3 | 157 | 14.80% | 1.10 (1.01-1.20) | 0.032 | 1.09 (0.93-1.27) | 0.301 |
| MMSE | 161 | 14.30% | 0.99 (0.85-1.14) | 0.856 | 1.21 (0.92-1.60) | 0.177 |
| MoCA | 114 | 11.80% | 0.81 (0.49-1.33) | 0.399 | 0.41 (0.13-1.29) | 0.129 |

PD- Parkinson's disease; UPDRS3- Unified Parkinson's Disease Rating Scale Part 3; MMSE- Mini Mental Stata Examination; MoCA- Montreal Cognitive Assessment.

There was some evidence for a causal effect of liability towards T2DM on PD progression measured using part 3 of the UPDRS (IVW OR 1.10, 95% CI 1.01-1.10; p=0.032), but no convincing evidence for progression in the MoCA (IVW OR 0.81, 95% CI 0.49-1.33; p=0.399) or MMSE scores (IVW OR=0.99, 95% CI 0.85-1.14; p=0.856). The directions of effect from all MR analyses were consistent with the effects from the observational study meta-analyses. Power calculations for the three progression markers suggested 12-15% statistical power to detect an effect, which suggests that progression analyses in particular may have been severely underpowered. We did not detect evidence of significant heterogeneity or bias arising from pleiotropic SNPs, utilizing a variety of sensitivity tests.

## DISCUSSION

In the present study, we used meta-analysis of observational and genetic data to investigate the role of T2DM as a determinant of both PD risk and progression. Results from the observational data meta-analyses and MR analyses were generally concordant; there was evidence from both methods that T2DM increases the risk of future PD. Our results also show that T2DM may also increase the rate of motor progression of PD, with weaker support for an effect on cognitive progression. However overall, treatment and/or prevention of T2DM may lead to a reduction in new diagnoses of PD and slowing of the progression of PD.

The effect of T2DM on PD risk was clearest in the highest quality prospective studies, which in turn should be less affected by observer and selection bias. Unlike previous meta-analyses looking at the association between diabetes and PD, we specifically looked at T2DM for the main risk meta-analysis.^4,5,6^ When we relaxed the exposure definition to include all studies of diabetes, an overall null association between diabetes and risk of PD was found. We also observed in both risk analyses a now-recognised phenomenon of divergence in pooled effect estimates by study design, such that case-control studies tend to be associated with lower risk (even inverse risk) of PD and cohort studies with increased risk of PD.^2,4,5^ This divergence appeared to be greater with lower quality studies and the phenomenon is discussed further below.

We extended observations about the effect of T2DM on PD risk to evaluate the limited available data on whether diabetes also affects PD progression. In data from observation studies, we observed evidence for an association between diabetes and PD progression on both motor and cognitive scales. However, evidence for this being causal was only observed for an effect on PD motor scores in the MR part of the analysis, whereas there was an absence of evidence for an effect on PD cognitive progression. These progression MR analyses lacked statistical power relative to the risk MR analysis and all observational study analyses.

Alongside epidemiological evidence, there is increasing evidence for shared biology between T2DM and PD. In T2DM, pancreatic islet amyloid polypeptide (IAPP) or amylin, aggregates to form amyloid plaques in pancreatic cells.^24^ Similarly, PD is pathologically defined by accumulation of alpha-synuclein intra-neuronally. There is some evidence to suggest that alpha-synuclein aggregation in PD occurs faster in the presence of IAPP.^25^ Whilst circulating insulin may have a neuroprotective role, systemic and local insulin resistance can influence pathways known to be important in PD pathogenesis, including those that relate to mitochondrial dysfunction, neuro-inflammation and synaptic plasticity.^1, 2^ Dopamine uptake is enhanced in the presence of insulin and dopamine release via enhanced cholinergic interneuron excitability resulting in the activation of nicotinic acetylcholine receptors.^36^ The links between T2DM and neurodegeneration exist not only with PD. The AKT pathway is one of several insulin signalling pathways and its overactivation has been linked to aggravation of Alzheimer’s disease pathology too.^26^ Similarly, in PD, the AKT pathway is altered causing an overexpression of GSK-3β which enhances the formation of neurofibrillary tangles and contributes to PD dementia.^37^ As the global burden of T2DM rises sharply, it becomes increasingly important to understand its potential role in neurodegeneration.^27^

Returning to the observation that associations between diabetes and PD diverge by study design, one explanation for this is survivor bias. Diabetes is associated with premature mortality and so an inverse association between diabetes and PD risk may be a result of greater mid-life mortality in patients with diabetes.^6^ Survivor bias may occur in case-control settings, but also in prospective cohorts in which the exposure occurs prior to the period of observation.^35^ We explored the potential role of survivor bias by examining case-control and cohort studies separately (discussed above), the effect of average age of participants in studies, the gender ratio, and finally by considering cohort studies separately depending on when the period of observation started relative to the onset of the exposure (T2DM). Average age and gender ratio had no major bearing on observational study estimates, but when cohort studies were separated according to the period of observation relative to the onset of the exposure, this yielded potentially revealing findings. For one of the cohort studies, the onset of T2DM was prior to the period of observation and this suggested a negative association with risk of PD but wide confidence intervals that crossed the null. Two of the cohort studies enrolled incident cases of T2DM and the pooled effect was a precise estimate in favour of increased risk of PD. The four remaining cohort studies used mixed exposure ascertainment (self-reported history of diabetes at enrolment i.e. *unobserved*, and new incident cases i.e. *observed*). The pooled point estimate from these studies was similar to the point estimate when the exposure was fully observed, but the confidence intervals were much wider. Overall these sub-analyses indicated that only the highest quality studies tend to support the association between T2DM and PD risk, whereas poorer quality studies and those that are prone to the influence of survivor bias may under-estimate it.

Strengths of this study are the specific focus on the role of T2DM for most of the analyses, as well as the use of observational and genetic data to draw out causal inferences about the effect on PD risk and progression. We used several sub-analyses to further investigate the nature of associations which we observed. Limitations include the possibility of bias (discussed above), generalizability, limited statistical power for the progression MR analyses, and being unable to take account of the effect of treatment of T2DM and severity.

When it comes to generalizability, an important consideration is that the majority of the observational data and all of the genetic data were derived from patients of European ancestry. Hence, the results cannot be readily generalized to all populations. People of South-Asian and African-Caribbean descent are at greatest risk of T2DM.^28^ The effect of T2DM on motor and cognitive progression of PD may differ by ethnicity, as a result of currently unknown but population-specific genetic and comorbid determinants and interactions. In the setting of dementia, one cohort study investigated whether diabetes was associated with a greater risk of mild cognitive impairment (MCI) in different ethnic groups.^29^ This study revealed that the prevalence of diabetes was highest in Hispanics (30%), and the attributable risk of MCI due to T2DM was 11% in Hispanics compared to 4.6% in non-Hispanics.^29^ It is possible that the prevalence of diabetes in different ethnic groups may in part account for differences in PD prevalence and phenotype.^30^

The findings from this study do not consider the effect of anti-diabetic drugs on PD risk and progression. The repurposing of drugs used to treat T2DM for PD has been a major driver of the interest between the two conditions. In a double-blind, placebo-controlled trial, it was shown that the GLP-1 analogue Exenatide, may have an effect on reducing PD severity after a prolonged washout period, raising the possibility of a disease-modifying effect.^31^ A recent cohort study from the same group reported evidence that patients with T2DM taking certain classes of drugs (GLP-1 receptor antagonists and DPP4 inhibitors) were at lower risk of PD than patients taking other oral antidiabetic drugs.^32^ The same study reported on the association between T2DM with PD (RR 2.65, 95% CI 2.58-2.71), but this was cross-sectional and was not included in our main analysis.^32^

Finally, our analysis does not take account of the likely important role of T2DM severity. T2DM represents a continuum ranging from those that are unaware of their disease state, to those that are aware but do not comply with management and are uncontrolled, as well as those that are diet-controlled, tablet-controlled or insulin-controlled. This spectrum of T2DM was not accounted for in our analyses, although biomarkers of glycaemic control are well established and may be used in future studies to further study the effects of T2DM severity on PD.^33^ In support of this, a recent study reported evidence in non-diabetic patients that midlife variability in glycaemic control was associated with future PD.^34^

In conclusion, we observe convincing evidence from the highest quality observational studies that T2DM is associated with an increased risk of PD, as well as some evidence that it may contribute to faster PD motor and cognitive progression. More studies are needed to explore the role of T2DM as a determinant of PD progression, as well as the strategies to modify this effect. Future studies, both observational and genetic, should seek to include greater representation of minority ethnic groups, many of whom experience a greater burden of T2DM. Also, treating T2DM may slow down the progression of PD; thus, a careful screening for T2DM and early treatment of T2DM in PD patients may be advisable. Finally, survival bias may be an important contributor to inverse associations between risk factors that cause premature mortality and age-related conditions, and this possibility warrants specific study.

## Data Availability

All data is available from the studies used.

## Notes

### Competing Interest Statement

ZGO received consultancy fees from Lysosomal Therapeutics Inc. (LTI), Idorsia, Prevail Therapeutics, Inceptions Sciences (now Ventus), Ono Therapeutics, Neuron23, Handl Therapeutics, Denali and Deerfield. Dr Noyce reports grants from the Barts Charity, Parkinson's UK, Aligning Science Across Parkinson's and Michael J Fox Foundation, and the Virginia Kieley Benefaction. Personal fees/honoraria from Britannia, BIAL, AbbVie, Global Kinetics Corporation, Profile, Biogen, Roche and UCB, outside of the submitted work.

