## Supplementary table 1 for "Type 2 diabetes as a determinant of Parkinson’s disease risk and progression"

Supplementary table 1: Studies included in the T2DM-specific meta-analysis from the literature review.

| Study | Study Design | Ethnicity of participants | Follow-Up (years) | Average Age (years) | Sample Size | Effect Estimate (95% Confidence Interval) | Quality Score |
| --- | --- | --- | --- | --- | --- | --- | --- |
| Herishanu et al. 2001 | Case-Control | Middle-Eastern |  |  | 93 case and 93 controls | 0.35 (0.15, 0.75) | Poor |
| Powers et al. 2006 | Case-Control | Caucasian |  | 70 | 352 cases and 484 controls | 0.62 (0.38, 1.01) | Fair |
| Palacios et al. 2011 | Cohort | Caucasian | 13 | 71.6 | 656 | 0.88 (0.62, 1.25) | Good |
| Yang et al. 2017 | Cohort | East-Asian | 7.3 | 56 | 36,294 (T2DM patients); 108,882 (non-T2DM patients) | 1.19 (1.08, 1.32) | Good |
| Jacobs et al. 2020 | Cohort | Caucasian | 12 | 62.7 | 501,682 | 1.27 (1.03, 1.57) | Good |
| De Pablo-Fernandez et al. 2018 | Cohort | Caucasian |  | 50 | 2,017,115 (T2DM cohort); 7,173,208 (reference cohort) | 1.32 (1.29, 1.35) | Good |
| Driver et al. 2008 | Cohort | Caucasian | 23.1 | 73.1 | 21,841 | 1.34 (1.01, 1.77) | Good |
| Xu et al. 2011 | Cohort | Caucasian |  | 66.7 | 1,565 | 1.41 (1.2, 1.66) | Fair |
| Hu et al. 2007 | Cohort | Caucasian | 18 | 48.8 | 51,552 | 1.83 (1.21, 2.76) | Good |

^[1]^

1. T2DM- type 2 diabetes mellitus.
