## Supplementary table 2 for "Type 2 diabetes as a determinant of Parkinson’s disease risk and progression"

Supplementary table 2: Studies included in the any diabetes meta-analysis from the literature review.

| Study | Study Design | Ethnicity of participants | Follow-up (years) | Average Age | Sample Size | Effect Estimate (95% Confidence Interval) | Quality Score |
| --- | --- | --- | --- | --- | --- | --- | --- |
| Miyake et al. 2010 | Case-Control | East Asian |  | 67.7 | 249 cases and 368 controls | 0.38 (0.17, 0.79) | Poor |
| D’Amelio et al. 2009 | Case-Control | Caucasian |  | 66.7 | 318 cases, 318 controls | 0.4 (0.2, 0.8) | Good |
| Kessler 1972 | Case-Control | Caucasian |  | 67.8 | 228 cases, 228 controls | 0.58 (0.3, 1.1) | Poor |
| Savica et al. 2012 | Case-Control | Caucasian |  | 71 | 202 cases, 202 controls | 0.67 (0.31, 1.48) | Fair |
| Rugbjerg et al. 2009 | Case-Control | Caucasian |  | 73 | 13,695 cases, 68445 controls | 1.10 (0.8, 1.5) | Fair |
| Schernhammer et al. 2011 | Case-Control | Caucasian |  | 72.2 | 1,931 cases, 9,651 controls | 1.35 (1.1, 1.65) | Fair |
| Morano et al. 1994 | Case-Control | Caucasian |  | 68.2 | 74 cases and 148 controls | 1.39 (0.63, 3.05) | Poor |
| Leibson et al. 2006 | Cohort | Caucasian |  | 70 | 202 (PD patients), 202 (reference cohort) | 0.70 (0.4, 1.4) | Poor |
| Simon et al. 2007 | Cohort | Caucasian | 22.9 | 66.6 | 171,879 | 1.04 (0.74, 1.46) | Fair |
| Grandinettei et al. 1994 | Cohort | Caucasian | 26 | 69.7 | 8,006 | 1.20 (0.67, 2.12) | Poor |
| Kim et al. 2018 | Cohort | East Asian | 10 | 64.5 | 7,746 | 1.26 (1.19, 1.33) | Poor |
| Sun et al. 2012 | Cohort | East Asian |  |  | 603,416 (diabetic patients); 472,188 (non-diabetic cohort) | 1.61 (1.56, 1.66) | Good |
| Skeie et al. 2013 | Cohort | Caucasian |  | 67.3 | 212 (PD cohort), 175 (control cohort) | 1.94 (0.82, 4.57) | Fair |
| Becker et al. 2008 | Cross-Sectional | Caucasian |  |  | 3,637 cases, 3,637 controls | 0.95 (0.8, 1.14) | Good |
| De Pablo-Fernandez et al. 2017 | Cross-Sectional | Caucasian |  | 73 | 79 cases, 4919 controls | 0.19 (0.9, 3.98) | Poor |

^[1]^

1. PD- Parkinson's disease.
