## Supplementary table 3 for "Type 2 diabetes as a determinant of Parkinson’s disease risk and progression"

Supplementary table 3: Studies included in the progression meta-analysis from the literature review.

| Study | Study Design | Motor or Cognitive Progression | T2DM | Ethnicity of participants | Time Period (years) | Sample Size | SMD (95% Confidence Interval) | Quality Score |
| --- | --- | --- | --- | --- | --- | --- | --- | --- |
| Cereda et al. 2012 | Case-Control | Motor | Yes | Caucasian | 3 | 89 cases, 89 controls | 0.35 (0.06, 0.65) | Fair |
| Malek et al. 2016 | Cohort | Motor | No | Caucasian | 3.5 | 1,759 | 0.62 (0.41, 0.83) | Good |
| Pagano et al. 2018 | Case-Control | Motor  Cognitive | Yes | Caucasian | 3 | 25 cases, 14 controls | 0.83 (0.21, 1.45)  -0.83 (-2.38, -0.08) | Good |
| Ong et al. 2017 | Cross-Sectional | Cognitive | No | Caucasian | 3 | 12 cases, 65 controls | -0.95 (-1.62, -0.27) | Fair |

^[1]^

1. T2DM- type 2 diabetes mellitus; SMD- standardised mean difference.
