## Supplementary table 4 for "Type 2 diabetes as a determinant of Parkinson’s disease risk and progression"

Supplementary table 4: Newcastle Ottawa Scale quality assessment of studies investigating the effect of T2DM on PD risk.

|  | Selection | | | | Comparability | Outcome | | | Quality Score |
| --- | --- | --- | --- | --- | --- | --- | --- | --- | --- |
| Study | Representativeness of exposed cohort | Selection of the non-exposed cohort from same source as exposed cohort | Ascertainment of exposure | Demonstration that outcome of interest was not present at start of study | Comparability of cohorts on the basis of the design or analysis controlled for confounders | Assessment of outcome | Sufficient follow-up | Adequacy of follow up cohorts |  |
| De Pablo-Fernandez et al. 2018 | Participants were truly representative of patients with T2DM and were excluded if they had PD. | Yes | Secure record- ICD-10 code E11 (diagnosed with T2DM) from the English National Hospital Episode Statistics | Yes | Sex, calendar year of cohort entry, age, region of residence, and quintile Index of Multiple Deprivation score of patients. | Record-linkage | Follow up not specified | No statement | Good |
| Hu et al. 2007 | Participants were truly representative of patients with T2DM in Finland. 5 geographic areas of Finland were covered. | Yes | Self-report questionnaire | Yes | Age, sex, study year, BMI, systolic BP, cholesterol, education, alcohol consumption, tea consumption, coffee consumption, cigarette smoking, leisure-time physical activity and education. | All patients diagnosed with PD according to the criteria set by the Institution, the diagnosis is based on medical history, clinical examination. The diagnosis needs to be done by a consultant. | Yes- mean follow up of 18 years. | Complete follow-up of all the patients | Good |
| Xu et al. 2011 | Participants were truly representative of patients with T2DM via the National Institutes of Health-AARP Diet and Healthy Study | Yes | Self-report | Yes | Baseline age, race, sec, smoking status, education, physical activity and BMI. | Diagnosed by Doctor | Not specified | No statement | Fair |
| Driver et al. 2008 | Somewhat representative because females with PD were not included. | Yes | Self-report questionnaire | Yes | Age and smoking status | Self-report questionnaire | Yes Mean-23.1 years | Complete follow-up of all the patients | Good |
| Yang et al. 2017 | Participants were truly representative of patients with T2DM in Taiwan. Patients were found. | Yes | Taiwan’s National Health Research Institutes Dataset | Yes | Age and comorbidities | Record-linkage | Yes Mean-7.3 years | Complete follow-up of all the patients | Good |
| Palacios et al. 2011 | Participants were somewhat representative of patients with T2DM. | Yes | Self-report | Yes | Age smoking, education, BMI, physical activity, caloric intake, caffeine intake, pesticide exposure, alcohol intake and diary intake. | Neurologists contacted and medical records checked. | Yes Mean- 13 years | No statement | Good |
| Powers et al. 2006 | Participants were truly representative of patients with PD. | Yes | The Group Health Cooperative Health Maintenance Organisation. | Yes | Age, smoking, education and ethnicity | Diagnosed by neurologists | Follow up not specified | No statement | Fair |
| Herishanu et al. 2001 | Participants were truly representative of patients with PD. | Yes | Outpatient PD clinical of Soroka University Medical Centre. | Yes | No description | No description | Follow up not specified | No statement | Poor |
| Jacobs et al. 2020 | Participants were truly representative of patients with PD. | Yes | Linked Hospital Episode Statistics ICD codes or self-report | No | Age, sex, Townsend deprivation index at recruitment and ethnicity. | No description | Yes Mean- 12 years | Complete follow-up of all the patients | Good |

^[1]^

1. T2DM- type 2 diabetes mellitus, PD- Parkinson's disease; ICD-10- International Classification of Disease, Tenth Revision; BMI- body mass index; BP- blood pressure;
