## Supplementary table 5 for "Type 2 diabetes as a determinant of Parkinson’s disease risk and progression"

Supplementary table 5: Newcastle Ottawa Scale quality assessment of studies investigating the effect of any diabetes on PD risk.

|  | Selection | | | | Comparability | Outcome | | | Quality Score |
| --- | --- | --- | --- | --- | --- | --- | --- | --- | --- |
| Study | Representative-ness of exposed cohort | Selection of the non-exposed cohort from same source as exposed cohort | Ascertainment of exposure | Demonstration that outcome of interest was not present at start of study | Comparability of cohorts on the basis of the design or analysis controlled for confounders | Assessment of outcome | Was follow-up long enough for outcomes to occur | Adequacy of follow up cohorts |  |
| Miyake et al. 2010 | Participants were truly representative of patients with DM in Japan. | Yes | Hospital | Yes | Sex, age, region of residence, pack-years of smoking, years of education, leisure-time exercise, BMI, dietary intake of energy, cholesterol, vitamin E, alcohol, coffee and the dietary glycaemic index. | Self-reporting Questionnaire | Follow up not specified | No statement | Poor |
| Leibson et al. 2006 | Participants were truly representative of patients with PD. | Yes | Census | No | Not specified | No description | Not specified | Not specified | Poor |
| Skeie et al. 2013 | Participants were truly representative of patients with PD. | Yes | Norwegian PakWest study | Yes | Age | Structured interview | Not specified | Not specified | Fair |
| Morano et al. 1994 | Participants are somewhat of patients with PD. | Yes | Hospitals | No | Not specified | Not specified | Not specified | Not specified | Poor |
| Savica et al. 2012 | Participants were truly representative of patients with PD | No | Rochester Epidemiology Project | Yes | Age, sex, cigarette smoking and coffee consumption | Record-linkage | Not specified | Not specified | Fair |
| Rugbjerg et al. 2009 | Participants were truly representative of patients with PD in Denmark | No | Danish National Hospital Register | Yes | Chronic obstructive pulmonary disease and sex | Hospital register | Not specified | Not specified | Fair |
| Kessler 1972 | Participants were truly representative of patients with PD | Yes | Commercial sources | No | Age | Structured interview | Not specified | Note specified | Poor |
| Grandinettei et al. 1994 | Participants were somewhat of patients with PD | No | Medical records | No | Age | Not specified | Yes- 26 years | Not specified | Poor |
| Kim et al. 2018 | Participants were truly representative of patients with PD in South Korea | Yes | National Health Insurance Database | No | Not specified | Health insurance claims | Yes- 10 years | Not specified | Poor |
| Schernhammer et al. 2011 | Participants were truly representative of patients with PD | No | Danish Hospital Register | Yes | Age, sex and chronic obstructive pulmonary disease | Danish Hospital Register | Not specified | Not specified | Fair |
| De Pablo-Fernandez et al. 2017 | Participants were truly representative of patients with PD | Yes | NEDICES study | Yes | Age, sex, hypertension, dyslipidaemia, antidiabetic treatment, alcohol consumption, smoking status, BMI, presence of cerebrovascular disease and treatment with potential parkinsonism-inducing drugs. | Self-report | Not specified | Not specified | Poor |
| Simon et al. 2007 | Participants were somewhat representative of patients with PD | No | Nurses’ Health Study | No | Age and smoking status | Self-reported history | Yes- 22.9 years | Not specified | Fair |
| Becker at al., 2008 | Participants were truly representative of patients with PD | Yes | UK- based General Practice Research Database | Yes | BMI, smoking, asthma/COPD, dementia, hypertension, ischemic heart disease, congestive heart failure, stroke/transient ischemic attack, arrhythmia, hyperlipidaemia, epilepsy, affective disorders, schizophrenia, and neurotic and somatoform disorders. | Patient records | Not specified | Not specified | Good |
| D'Amelio et al. 2009 | Participants were truly representative of patients with PD in Italy | Yes | Neurological Department of Palermo | No | BMI, smoking habit, education and occupational status | Semi-structured questionnaire | Not specified | Subjects lost to follow up unlikely to introduce bias- number lost less than 20% | Good |
| Sun et al. 2012 | Participants were truly representative of patients with DM in Taiwan | Yes | NHI claim data of Taiwan | Yes | Age, sex, geographic area, urbanisation status, hypertension, hyperlipidaemia and cardiovascular disease. | Hospital records | Yes- 1 year | Not specified | Good |

^[1]^

1. PD- Parkinson's disease; T2DM- type 2 diabetes mellitus; BMI- body mass index; COPD- chronic obstructive pulmonary disease; DM- disease mellitus.
