## Supplementary table 6 for "Type 2 diabetes as a determinant of Parkinson’s disease risk and progression"

Supplementary table 6: Newcastle Ottawa Scale quality assessment of studies investigating the effect of diabetes on PD progression.

|  | Selection | | | | Comparability | Outcome | | | Quality Score |
| --- | --- | --- | --- | --- | --- | --- | --- | --- | --- |
| Study | Representativeness of exposed cohort | Selection of the non-exposed cohort from same source as exposed cohort | Ascertainment of exposure | Demonstration that outcome of interest was not present at start of study | Comparability of cohorts on the basis of the design or analysis controlled for confounders | Assessment of outcome | Sufficient follow-up | Adequacy of follow up cohorts |  |
| Cereda et al. 2012 | Participants were truly representative of patients with PD and T2DM. | Yes | The Parkinson Institute research database | Yes | No description | UPDRS scale | Yes- mean 3 years. | Subjects lost to follow up unlikely to introduce bias- number lost less than 20% | Fair |
| Malek et al. 2016 | Participants were truly representative of patients with PD and T2DM. | Yes | Tracking Parkinson’s study | Yes | All vascular risk factors | UPDRS scale | Yes- mean 2.6 years. | Subjects lost to follow up unlikely to introduce bias- number lost less than 20% | Good |
| Pagano et al. 2018 | Participants were truly representative of patients with PD and T2DM. | Yes | Parkinson's Progression Markers Initiative database | Yes | Sex, age, H&Y stage and MDS-UPDRS Part III score | UPDRS scale and MoCA | Yes- 3 years. | No statement | Good |
| Ong et al. 2017 | Participants were truly representative of patients with PD and T2DM. | Yes | No description | Yes | No description | MoCA | Yes- 36 months | No statement | Fair^[1]1^ |

1. UPDRS- Unified Parkinson's Rating Scale; MoCA- Montreal Cognitive Asessment
