## Supplementary table 7 for "Type 2 diabetes as a determinant of Parkinson’s disease risk and progression"

Supplementary table 7: MR analysis between exposure (T2DM) and outcomes (PD-risk and progression).

| Outcome | N, SNPs | Power | Inverse variance weighted | | | | MR Egger | | | | Weighted median | | | |
| --- | --- | --- | --- | --- | --- | --- | --- | --- | --- | --- | --- | --- | --- | --- |
|  |  |  | OR (95% CI) | | | p-value | OR (95% CI) | | | p-value | OR (95% CI) | | | p-value |
| PD risk | 185 | 38.00% | 1.08 (1.02-1.14) | | | 0.010 | 1.05 (0.93-1.17) | | | 0.427 | 1.05 (0.96-1.16) | | | 0.299 |
| Continuous PD progression traits | | | | | | | | | | | | | | |
| UPDRS3 | 157 | 14.80% | 1.10 (1.01-1.20) | | | 0.032 | 1.09 (0.93-1.27) | | | 0.301 | 1.14 (0.98-1.33) | | | 0.101 |
| MMSE | 161 | 14.30% | 0.99 (0.85-1.14) | | | 0.856 | 1.21 (0.92-1.60) | | | 0.177 | 1.02 (0.77-1.35) | | | 0.897 |
| MoCA | 114 | 11.80% | 0.81 (0.49-1.33) | | | 0.399 | 0.41 (0.13-1.29) | | | 0.129 | 0.71 (0.33-1.51) | | | 0.371 |
| Outcome | Heterogeneity tests | | | | | | Test for directional horizontal pleiotropy | | | |  |  |  |  |
|  | MR Egger | | | Inverse variance weighted | | | Egger_intercept | SE | p-value | MR-PRESSO global |  |  |  |  |
|  | Q | Q_df | Q_pval | Q | Q_df | Q_pval |  |  |  | pval |  |  |  |  |
| PD risk | 126.369 | 183.000 | 1.00 | 126.73 | 184 | 1.00 | 0.002 | 0.004 | 0.550 | 0.990 |  |  |  |  |
| Continuous PD progression traits | | | | | | | | |  |  |  |  |  |  |
| UPDRS3 | 140.270 | 156 | 0.81 | 140.29 | 157 | 0.83 | 0.001 | 0.005 | 0.885 | 0.841 |  |  |  |  |
| MMSE | 135.158 | 159 | 0.92 | 138.10 | 160 | 0.89 | -0.015 | 0.009 | 0.088 | 0.848 |  |  |  |  |
| MoCA | 124.287 | 112 | 0.20 | 126.13 | 113 | 0.19 | 0.045 | 0.035 | 0.200 | 0.233 |  |  |  |  |

^[1]^

1. PD- Parkinson's disease; SE- standard error; UPDRS3- Unified Parkinson's Disease Rating Scale Part 3; MMSE- Mini Mental Stata Examination; MoCA- Montreal Cognitive Assessment.
