## Supplementary figures and images for "Type 2 diabetes as a determinant of Parkinson’s disease risk and progression"

### Supplementary figure 1

Supplementary Figure 1: Funnel plot generated for T2DM-specific studies.

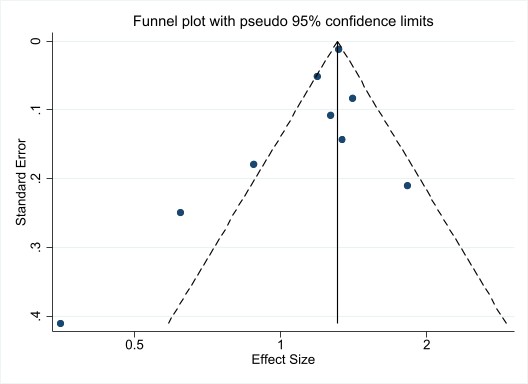

### Supplementary figure 3

Supplementary figure 3: An observed exposure (T2DM) increases the risk of PD.

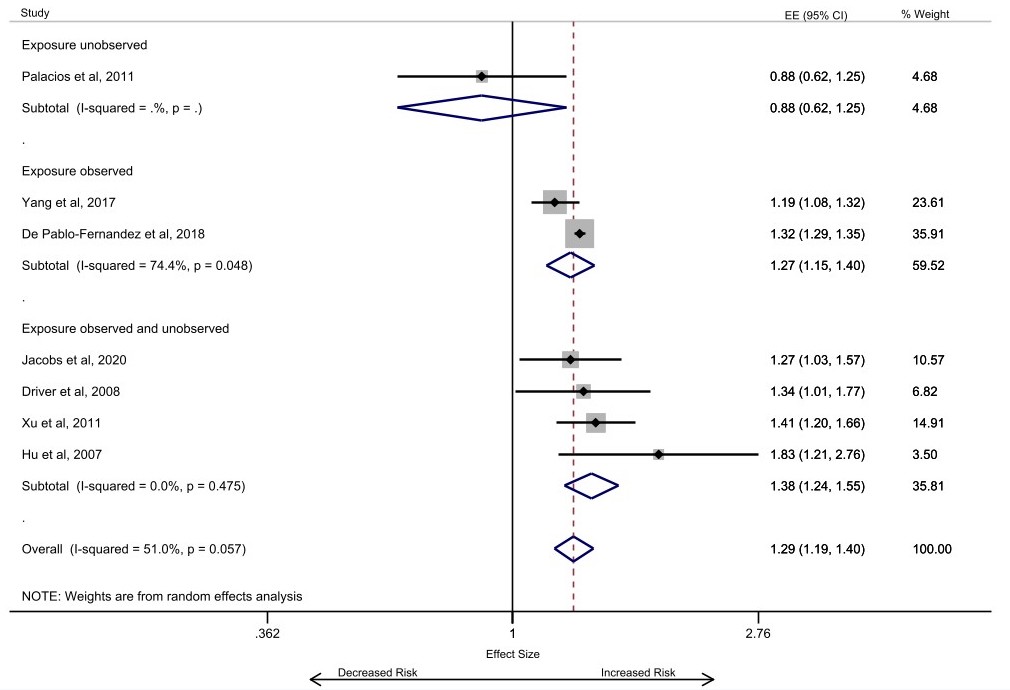
