## Supplementary figure 2 for "Type 2 diabetes as a determinant of Parkinson’s disease risk and progression"

Supplementary Figure 2: PD risk decreases as average age of participants increases in T2DM-specific cohort studies.

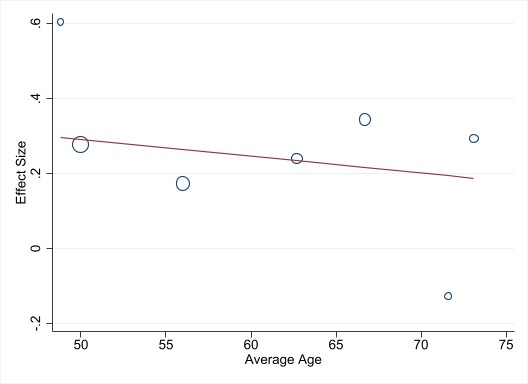
