## Supplementary figure 4 for "Type 2 diabetes as a determinant of Parkinson’s disease risk and progression"

Supplementary figure 4: Diabetes slightly increases the risk of Parkinson's disease (any diabetes).

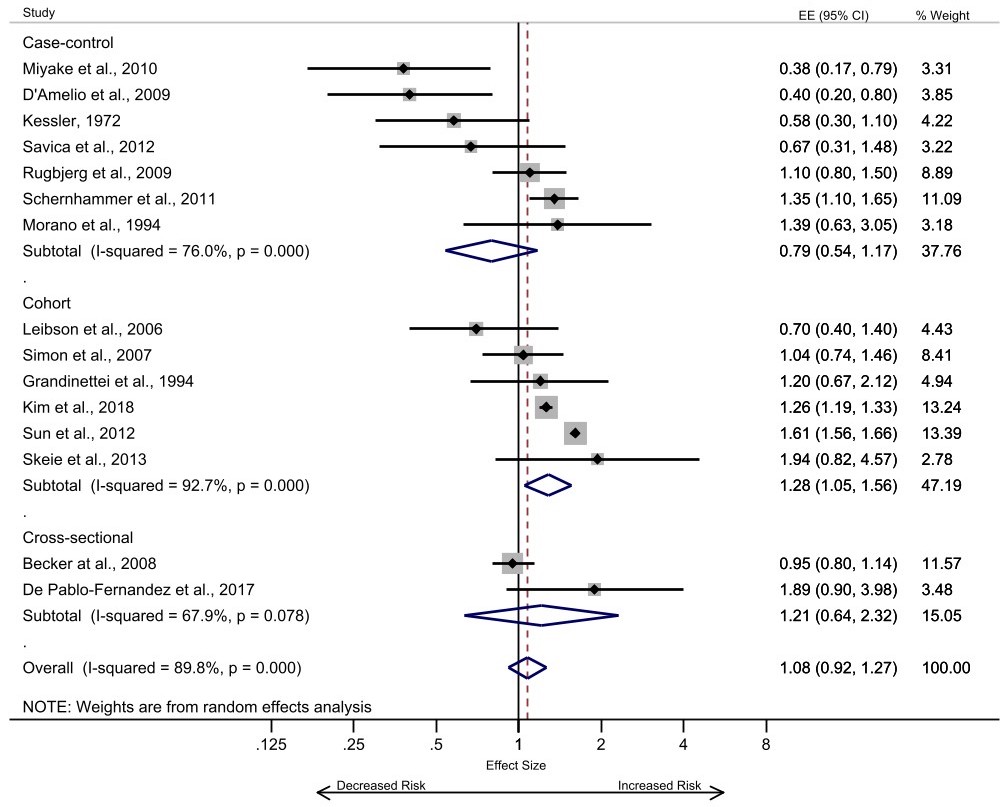
