## Supplementary figure 6 for "Type 2 diabetes as a determinant of Parkinson’s disease risk and progression"

Supplementary figure 6: Funnel plot generated after trim and fill analysis to account for publication bias. 1 study was imputed.

0

.1

.2

.3

.4

S

t

a

n

d

a

r

d

E

r

r

o

r

0.25

0.5

1

2

4

Effect Size

Funnel plot with pseudo 95% confidence limits
