## Supplementary figure 7 for "Type 2 diabetes as a determinant of Parkinson’s disease risk and progression"

Supplementary figure 7: PD risk increases as average age of participants increases in case-control studies (any diabetes).

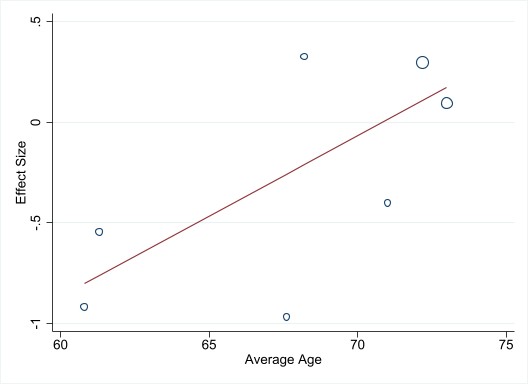
