## Supplementary figure 8 for "Type 2 diabetes as a determinant of Parkinson’s disease risk and progression"

Supplementary figure 8: PD risk decreases as the average age of participants increases in cohort studies (any diabetes).

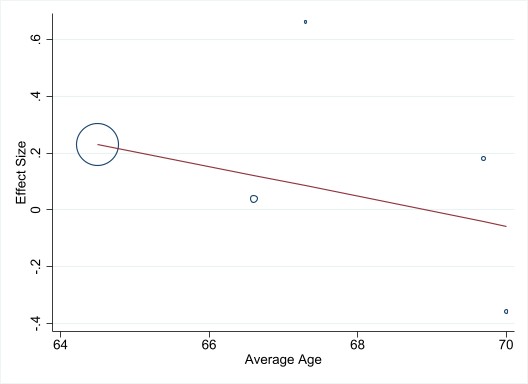
