## Supplementary figure 9 for "Type 2 diabetes as a determinant of Parkinson’s disease risk and progression"

Supplementary figure 9: In the pooled case-control studies, as the average age of the participants increases the PD risk increases (T2DM-specific and any diabetes studies).

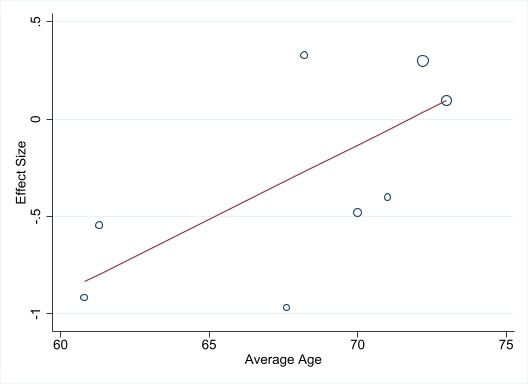
