## Supplementary figure 10 for "Type 2 diabetes as a determinant of Parkinson’s disease risk and progression"

Supplementary Figure 10: Forest plots showing point estimates of the exposures of interest; Diabetes as exposure. PD risk and progression as outcomes.

**PD risk as outcome**

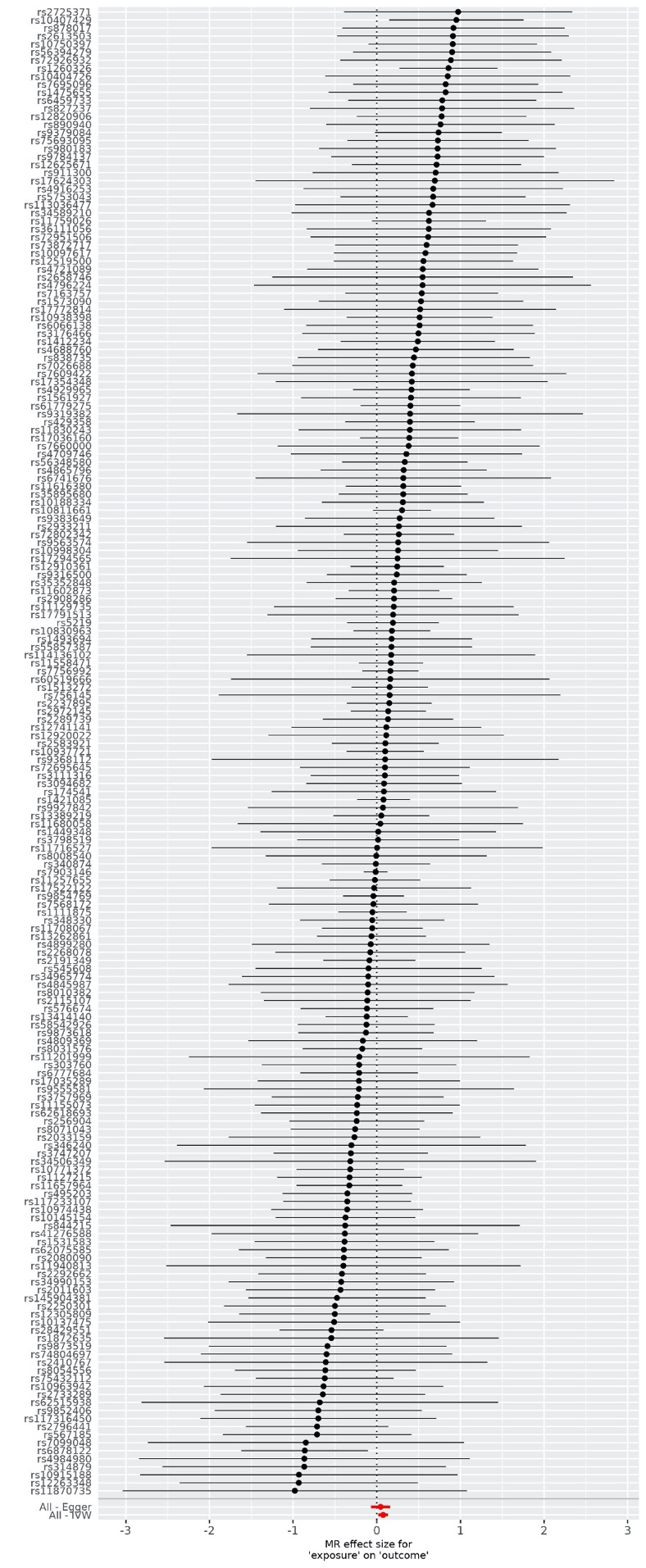

**MMSE in PD as outcome**

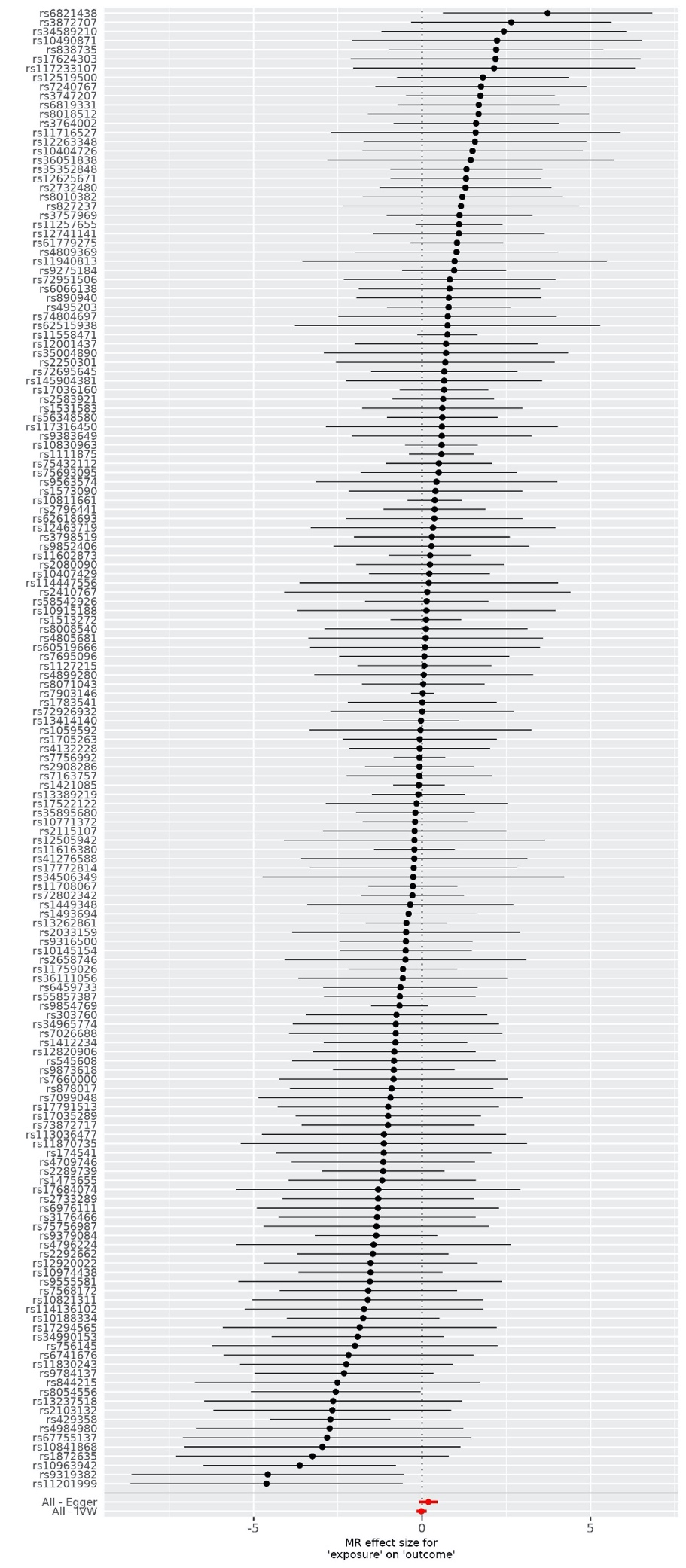

**MoCA in PD as outcome**

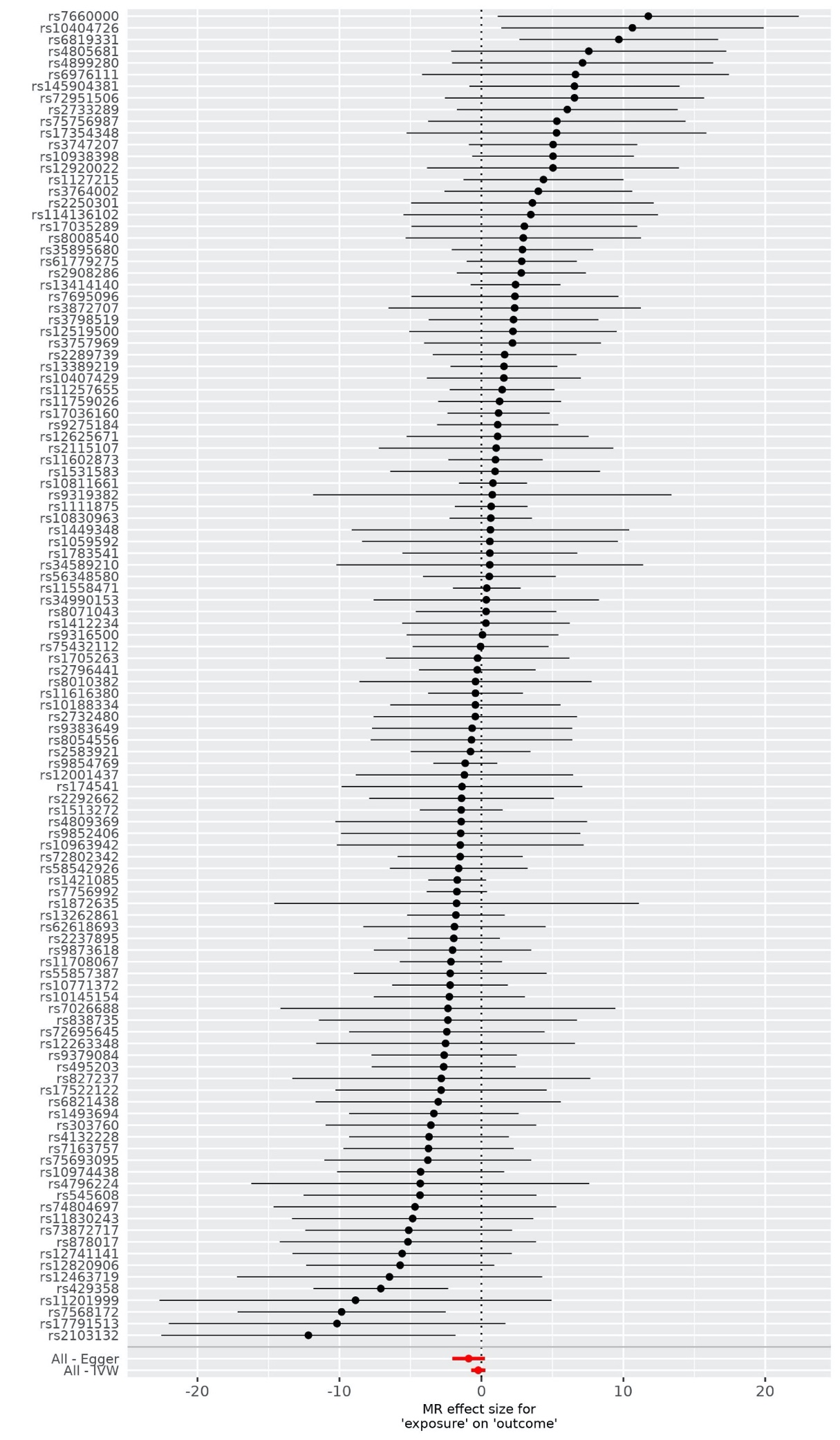

**UPDRS3 in PD as outcome**

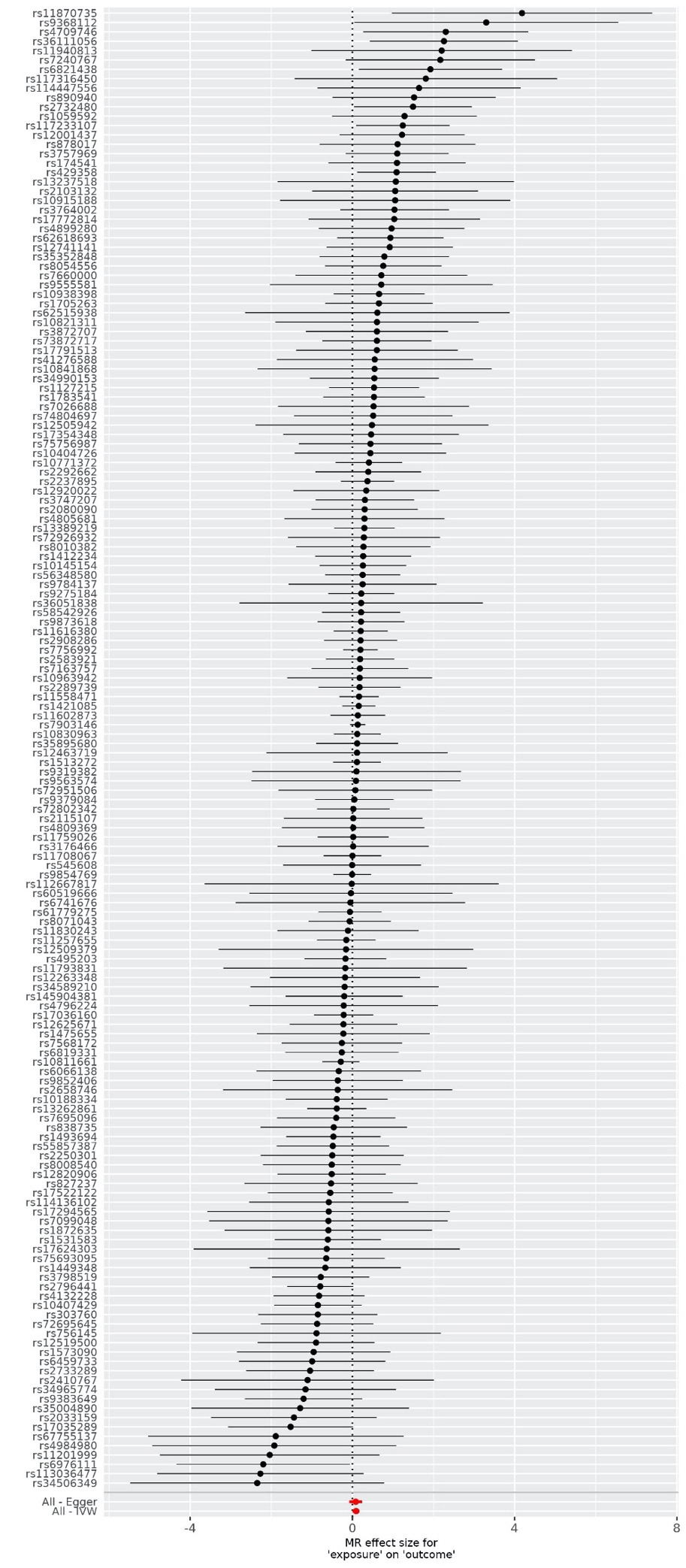
