## Supplementary figure 11 for "Type 2 diabetes as a determinant of Parkinson’s disease risk and progression"

Supplementary Figure 11: Funnel plots showing point estimates as the exposures of interest; Diabetes as exposure. PD risk and progression as outcomes.

**PD risk as outcome**

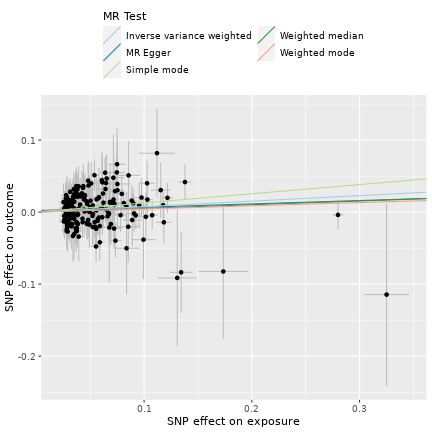

**MMSE in PD as outcome**
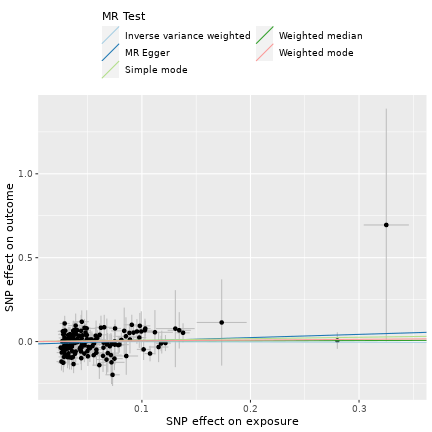

**MoCA in PD as outcome**
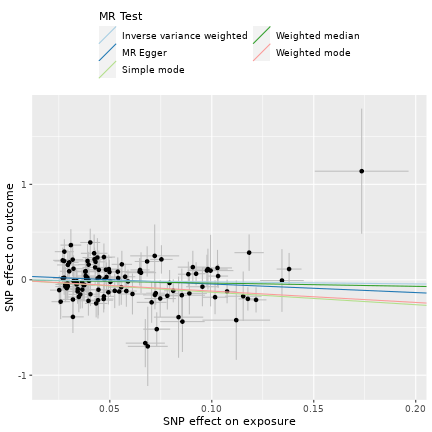

SEADL in PD as outcome

**UPDRS3 in PD as outcome**
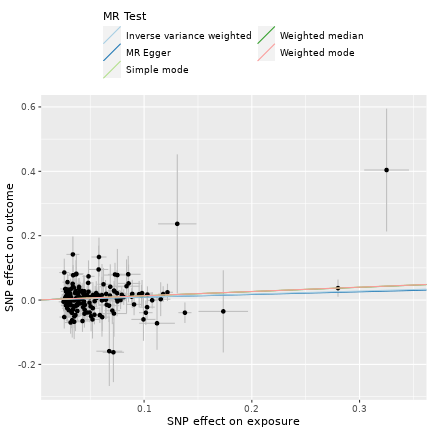
